# Effects Of Good Governance on Health System Performance: An empirical analysis of the WHO Europe region

**DOI:** 10.1101/2025.07.01.25330666

**Authors:** Levan Samadashvili, Gareth H Rees, Rosemary James, Alison Leary, Thomas Hughes-Waage, Cris Scotter

**Affiliations:** WHO consultant, Tbilisi, Georgia; ESAN University, Lima, Peru; Geneva Centre of Humanitarian Studies, Faculty of Medicine, University of Geneva, Geneva, Switzerland.; School of Health, London South Bank University, London, UK; WHO Consultant, London, UK; Royal College of Surgeons in Ireland (RCSI) Graduate School of Healthcare Management, Dublin, Ireland and World Health Organization Regional Office for Europe, Copenhagen, Denmark

**Keywords:** Health Governance, Health system performance, Performance measurement, Europe

## Abstract

**Context:** Governance in the health sector has gained more attention in the recent literature, although, the topic is still relatively under-examined. The WHO European Region comprises 53 member states that each have unique health systems, HWF planning and governance characteristics. These countries are struggling to develop optimal approaches to strengthen health systems and improve performance outcomes amidst local contexts, lack of financial resources, and varying supply and demand of health workers. While it has been widely recognized that a one-size-fits all approach cannot be successful and each country requires specific, context-bound solutions, it is also evident that there are certain key features and elements, or general enablers for a more successful health system performance. Good Governance should be regarded as one of such, and is possibly the most critical general enabler of a better functioning healthcare system.

**Methods:** To test this proposition, we use a set of universal indicators from publicly accessible data bases that were analysed in Microsoft Excel using its Pearsons correlation coefficient function for the sets of data pairs to identify an association between governance and health performance.

**Findings:** We found a positive association with variable strength between the variables’ pairs. We corroborated these findings with country clusters based on the other five WHO regions, where we also found the association found in Europe holds, also with some variation between clusters.

**Conclusions:** These results reveal that while linking governance to health system performance is a difficult task, there is an observable association that should be further studied.

**POLICY POINTS:**

- Governance is generally acknowledged as important for health system performance, though there is little empirical validation of this association.
- An association is confirmed in WHO Europe region countries in our study and by way of corroboration, in other WHO region country clusters.
- There is variation of the association country by country indicating that context matters and this requires further research.

## Introduction

Many countries are struggling to develop optimal approaches to strengthen health systems and improve outcomes amidst long waiting times for elective procedures, balancing resource and facility use, and increasing staff vacancies [1]. Health governance or the existence of strategic policy frameworks that are “combined with effective oversight, coalition-building, regulation, attention to system-design and accountability” [2, p.86] is recognized as an important contributor for health system outcomes. This is because the outcomes in health are affected by multiple factors outside the direct control of the health sector such as education, income, and individual living conditions [3]. Thus, governance encompasses directing a collection of practices that include “rules (both formal and informal), roles and responsibilities for collective action and decision-making in a system with diverse stakeholders” [4, p.2]. While it has been widely recognized that there cannot be one-size-fits all approach and each context requires specific tailored approaches [5], governance is considered to be “a practice” [6, p720] that is not only dependent on nationally determined settings, but is also determined through lower-levels of health system implementation. Thus, governance’s role and functions are increasingly included in multilevel frameworks used to assess health system functioning, which are constructed from certain key features, elements, or general enablers that contribute to health system performance [7]. While it is accepted that there are contextual and operationalization difficulties when seeking to conceptualize the measurement and effects of governance [6, 7, 8], it has been acknowledged that governance settings may impact the health workforce [9] and governmental influence [10].

As such, the accepted wisdom is that a health system with a set of ‘good’ governance practices should be a stronger health system or have better health system performance. However, while health sector governance continues to gain attention in the literature, clearly determining this accepted relationship has remained elusive and the role of governance in health system performance continues to be relatively under-examined or even neglected [6].

This article aims to add to the evidence base to bolster the accepted wisdom of a positive association between good governance and health system performance. We do this by analysing country-level governance and health system indicators for 52 of the WHO Europe region’s member states to determine any association and extend this analysis to the other WHO regions as form of corroboration. Thus, next we present more on the constructs of health governance and health system outcomes, their measurement frameworks and issues. We then present the data and methods that we used to detect an association. This is followed by our results, which we discuss, and close the article with a short conclusion.

### Health System Governance

Governance is the structure of decision making and coordination across a system or society and must be good enough to get things done or advance policies [11]. Its importance to health has been recognized for some time, with many of the seemingly intractable problems in global health suggested to be able to be addressed through improved global health governance [12]. It is also seen as a “recurrent problem” [11, p42], being indicated as a reason for policy failure, the reason for continuing the status quo and for a lack of change from ineptitude, conflicts of interest, bureaucratic rigidities or poorly thought-out policies [11]. COVID-19 has also revealed and reemphasized the importance of governance, particularly in the realm of international public health [13]. Hence, a key contributor to good policy and its implementation is to clearly identify these weaknesses of governance and gain a better understanding of them [11].

Significant to understanding governance in the health context is the distinction between ‘governance for health’, which includes the contribution of sectors outside of the sphere of health that impact health systems and ‘health governance’, which is the governance of the health system and strengthening its systems [14]. These definitional differences have some impacts given governance for health’s exogeneity of the determinants of health, as opposed the direct actions and derivation and initiating of polices that relate to health governance [15].

Governance is seen to be a core health system function broadly influencing all other functions, their intermediate outcomes and final goals. From this centrality to a health system’s performance has governance being included in a range of measurement frameworks [17]. Many of these frameworks tend to contain desirable attributes of governance and while not all frameworks contain the same attributes, there is considerable overlap [11]. These “handful of topics” [7, p47] are variously labelled as strategies, elements, principles, or functions depending on the frameworks’ authors perspectives.

Regardless of the naming variations, the health system frameworks aim to identifying the components of governance rather the actual components of good governance [11]. Importantly, there is no one-size-fits-all approach to good governance – each country must plan and implement in accordance with its unique context and resources.

Nonetheless, significant common ground can be found among the range of health system assessment frameworks and analyses, such as the five core functions of Health System Governance (HSG) [5], four governance sub-functions of the Health System Performance Assessment (HSPA) framework [7], the five governance domains of the TAPIC framework [11] and Barbazza, et al.’s eight sub-functions of governance [18]. Table 1 provides a summary of these health system assessment frameworks, revealing conceptual similarities between their constituent components, allowing the overlaps to be readily identified when placed together.

**Table 1:** Governance component overlaps in framework construction.

| **HSPA sub-functions** | **TAPIC domains** | **Core HSG Functions** | **Governance Sub-functions** |
| --- | --- | --- | --- |
| Policy and vision | Transparency | Policy formulation and strategic plans | Build partnerships |
| Stakeholder voice | Accountability | Intelligence | Formulate strategic direction |
| Information and intelligence | Participation | Regulation | Regulate |
| Legislation and regulation | Integrity | Collaboration and coalition | Establish organizational adequacy |
|  | Policy capacity | Accountability | Foster participation |
|  |  |  | Ensure accountability |
|  |  |  | Generate information |
|  |  |  | Promote transparency |

Source: [5, 7, 11, 18].

However, the existence of governance components in a framework does not necessarily guarantee optimal health system functionality. Some of these components may act to impede other virtuous system elements such as speed, efficiency, effectiveness, flexibility, creativity, empowerment and innovation [11].

Nevertheless, a number of health system studies have tended to substantiate the accepted wisdom of a positive relationship between good governance and health system performance. For example, Gostin and Mok found that the overall governance by the State that seemed to have more of an effect rather than how the health system is being operated and managed [12]. While other studies found particular components have been found to have led to the effect, for example, Debie et al.’s review found robust health intelligence and intersectoral collaboration and coalition as for effective pandemic response as well as identifying other components that presented challenges for achieving Universal Health Coverage (UHC) [5]. These studies conform to the general component categorization approach and point an influence of wider governance elements that may not be specifically health related to the overall quality of a country’s governance processes, such as dealing with corruption or ensuring that system and policy capability. As such, in a single country study Masefield et al. [19] found that political, structural, and financial challenges were barriers to effective health governance at all health system levels. However, not all studies have been so supportive. While echoing Macefield et al.’s findings on key mechanisms or governance components as contributors to health system outcomes, Ciccone et al.’s [20] synthesis of peer-reviewed literature found not all of the 30 studies in their review had positive associations between governance and health, leading them to conclude that governance by itself is not guaranteed to improve health and governance’s role and contribution deserves further research.

Moreover, the indicators that have been used to measure governance have been criticized, with detractors maintaining that these indicators are theoretical and biased, with a counter argument being that no better alternatives exist [21]. In response Andrews et al. argued that more appropriate governance indicators are required that have some theoretical grounding, a focus on specific engagement, emphasize outcomes and control for key contextual differences in comparing countries [21]. Another issue raised by Koller et al. for measuring governance is that of anti-corruption, transparency and accountability, which generally tend to be missing from efforts to promote UHC [22]. As such, the lack of a settled set of health governance measures and constructs has led to composite governance indices being developed, some that are based on existing indices such as the World Bank Worldwide Governance Indicators (WGI) [23] (see [15]).

### Heath System Performance

The literature in relation to health system performance or outcomes is similarly equivocal. Ciccone et al. [20] state that the empirical literature linking governance to health is relatively sparse, with the studies being diverse, exploring a single or particular governance mechanism and health outcome(s) within specific situations. These situations ranged from examining a particular process or structure with most, but not all, of the reviewed studies finding a positive association between governance and health. Similarly, Debie, et al.’s study found that the decentralization of health services to grass root levels, support of stakeholders, fair contribution and distribution of resources were key to achieving UHC and health security programmes [5]. Azimi, et al.’s [15] results on the other hand highlighted a unidirectional causality where governance was related to health system costs, which are represented by governmental health expenditures and patient health-related out-of-pocket expenses. However, Papanicolas et al. recommend that when assessing actual health system goals, the measures used should be expressed as outcomes rather than using health structure or process parameters. [17].

Thus, health system performance indices such as the UHC Service Coverage Index developed by WHO Global Health Observatory [24] and the Health Index Score developed by Legatum [25] are comprised of outcome-based measures to gauge levels of coverage and access to necessary services.

Such indices and their constituent data have been used to measure or compare UHC advancement (for example [26]), as well as to further develop simplified indices for health system performance assessment [27].

## Methods

Our data was sourced from indices reflecting governance and health system performance from publicly available global databases. The data sources included World Bank’s Worldwide Governance Indicator (WGI) for governance and the WHO UHC Service Coverage Index, the Legatum Health Index, and the Global Health Security (GHS) Index to operationalize health system performance measures.

We selected the variables for good governance, using three WGI components namely: regulatory quality, which captures the ability of the government to formulate and implement sound policies and regulations; government effectiveness, which captures perceptions of the quality of public services, the quality of the civil service and the degree of its independence from political pressures, the quality of policy formulation and implementation, and the credibility of the government’s commitment to such policies; and control for corruption, which captures perceptions of the extent to which public power is exercised for private gain across large and small forms of corruption, as well as “capture” of the state by elites and private interests [23]. For health system performance variables, we used the respective country scores from the Global Health Security Index (Economist Impact) [28], Universal Health Coverage (WHO), and Health Index Score (Legatum) (see Table 2).

**Table 2:** Governance and Health system performance data of the 52 WHO Europe region members states analysed in this study.

| WHO Europe Member States (n=52) | Global Health Security Index<sup>a</sup> | UHC Service Coverage Index<sup>b</sup> | Health Index Score<sup>c</sup> | WGI Government Effectiveness<sup>d</sup> | WGI Regulatory Quality<sup>d</sup> | WGI Control for Corruption<sup>d</sup> |
| --- | --- | --- | --- | --- | --- | --- |
| Albania | 45 | 64 | 73.9 | 50.95 | 58.57 | 31.90 |
| Andora* | 34.7 | 79 | n/a | 97.14 | 89.52 | 88.57 |
| Armenia | 61.8 | 68 | 73.5 | 40.00 | 57.14 | 57.62 |
| Austria | 56.9 | 85 | 80.2 | 93.81 | 87.14 | 85.71 |
| Azerbaijan | 34.7 | 66 | 71.2 | 58.57 | 50.48 | 21.43 |
| Belarus | 43.9 | 79 | 74.3 | 20.48 | 17.14 | 47.62 |
| Belgium | 59.3 | 86 | 80.6 | 83.33 | 86.67 | 89.52 |
| Bosnia and Herzegovina | 35.4 | 66 | 70.5 | 12.86 | 46.19 | 28.10 |
| Bulgaria | 59.9 | 73 | 74.1 | 45.71 | 66.67 | 48.10 |
| Croatia | 48.8 | 80 | 75.7 | 69.52 | 68.10 | 56.67 |
| Cyprus* | 41.9 | 81 | 79.2 | 73.81 | 76.67 | 63.33 |
| Czechia | 52.8 | 84 | 79.5 | 81.43 | 87.62 | 71.90 |
| Denmark | 64.4 | 82 | 81.1 | 98.57 | 97.62 | 10.00 |
| Estonia* | 55.5 | 79 | 77.7 | 88.57 | 92.86 | 90.00 |
| Finland | 70.9 | 86 | 81.2 | 98.10 | 99.05 | 99.52 |
| France | 61.9 | 85 | 80.5 | 85.24 | 85.71 | 89.05 |
| Georgia | 52.6 | 68 | 70.6 | 70.95 | 82.86 | 74.29 |
| Germany | 65.5 | 88 | 81.4 | 87.14 | 94.76 | 95.71 |
| Greece | 51.5 | 77 | 77.4 | 67.14 | 66.19 | 59.52 |
| Hungary | 54.4 | 79 | 76.7 | 70.48 | 67.62 | 55.24 |
| Iceland* | 48.5 | 89 | 82.7 | 94.76 | 91.90 | 95.24 |
| Ireland | 55.3 | 83 | 80 | 91.90 | 93.33 | 92.86 |
| Italy | 51.9 | 84 | 80.9 | 64.29 | 68.57 | 67.62 |
| Kazakhstan | 46.1 | 80 | 72.2 | 55.24 | 55.71 | 47.14 |
| Kyrgyzstan | 42.4 | 69 | 72.5 | 22.38 | 30.48 | 12.86 |
| Latvia* | 61.9 | 75 | 74.5 | 76.67 | 85.24 | 76.19 |
| Lithuania | 59.5 | 75 | 74.4 | 80.95 | 86.19 | 79.52 |
| Luxemburg* | 48.4 | 83 | 81.6 | 95.71 | 99.52 | 96.19 |
| Malta* | 40.2 | 85 | 80.5 | 77.14 | 74.29 | 62.86 |
| Moldova | 41 | 71 | 70.6 | 36.19 | 52.38 | 35.24 |
| Monaco* | 33.3 | 86 | n/a | 99.52 | 89.52 | 88.57 |
| Montenegro* | 44.1 | 72 | 68.4 | 51.43 | 65.71 | 52.86 |
| Netherlands | 64.7 | 85 | 82 | 96.19 | 96.19 | 97.14 |
| North Macedonia | 42.2 | 74 | 72.6 | 47.14 | 65.24 | 41.90 |
| Norway | 60.2 | 87 | 83 | 97.62 | 95.24 | 98.10 |
| Poland | 55.7 | 82 | 76.3 | 61.43 | 75.71 | 68.10 |
| Portugal | 54.7 | 88 | 77.4 | 80.48 | 73.33 | 77.14 |
| Romania | 45.7 | 78 | 73 | 46.19 | 61.90 | 51.43 |
| Russian Federation | 49.1 | 79 | 71.4 | 43.33 | 32.86 | 19.52 |
| San Marino* | 32.9 | 77 | n/a | 97.14 | 70.48 | 88.57 |
| Serbia | 45 | 72 | 71.9 | 52.86 | 53.33 | 35.71 |
| Slovakia | 54.4 | 82 | 76.7 | 68.57 | 77.62 | 60.00 |
| Slovenia* | 67.8 | 84 | 79.9 | 83.81 | 74.76 | 75.24 |
| Spain | 60.9 | 85 | 79.7 | 78.10 | 73.81 | 75.71 |
| Sweden | 64.9 | 85 | 82.3 | 95.24 | 96.67 | 97.62 |
| Switzerland | 29.3 | 86 | 82.1 | 99.05 | 95.71 | 96.67 |
| Tajikistan | 58.8 | 67 | 72.5 | 29.52 | 11.43 | 6.67 |
| Turkiye | 50 | 76 | 74.2 | 48.57 | 48.10 | 39.52 |
| Turkmenistan | 31.9 | 75 | 75.6 | 14.29 | 1.90 | 5.71 |
| Ukraine | 38.9 | 76 | 68.7 | 35.24 | 42.38 | 24.29 |
| United Kingdom | 67.2 | 88 | 78.3 | 86.19 | 90.95 | 93.33 |
| Uzbekistan | 39 | 75 | 76.3 | 40.48 | 31.90 | 22.38 |

Note: \* Identifies Small Country data set
Source: a2021 data
b WHO, 2021
c Legatum, 2023
d World Bank Data, 2021

These data were analysed in Microsoft Excel using its Pearsons correlation coefficient function for the sets of data pairs. The Pearsons correlation coefficient, commonly abbreviated to “*r*”, takes a value from −1 to +1, where a positive sign of the correlation coefficient reveals the existence of a positive correlation, while a negative sign reveals a negative correlation and a perfect correlation defined by a coefficient of 1 or −1 and a correlation coefficient of zero indicates that there is no linear association between the two variables [29]. These results rely a number of assumptions, for instance that data are derived from a random or representative sample, the variables are continuous, jointly normally distributed, random variables and the association, if one exists, is always linear between jointly normally distributed data [30]. Schober, Boer and Schwarte also caution the use of descriptors aligned to arbitrary and inconsistent ranges such as strong for example, moderate or weak, even though several categorizations with various transition points have been published [30]. Rather they suggest that “a specific coefficient should be interpreted as a measure of the strength of the relationship in the context of the posed scientific question” [30, p.1765]. Thus, we apply author cautions, by taking the accepted wisdom of the relationship between these two sets of variables to seek identify its strength and direction.

For the purposes of consistency, and for making sure that observed patterns are not due to coincidence, respective index scores were analysed at three different points in time. We also disaggregated the small countries of the region, as these are likely to share common characteristics in terms of political context, economic features, and constraints in resource capacity that maybe different to larger countries and from the pandemic have been found to have more of a dependency on larger neighbouring countries for trade, access to medicines and vaccines, and the shortage of and strain on health workers [32].

As a confirmation measure, we collected available data from the source databases from countries located in the five other WHO regions [33], compiled it in Excel and applied the Pearsons correlation coefficient function on these data.

## Results

The results from the 52 WHO Europe Region Countries are contained in Table 3, which presents the total country correlation scores, along with a grouping of countries with small countries taken out and then the correlation scores for the small country group. Figure 1 provides the graphical spread and line of best fit for each of the governance variables and the UHC data.

**Table 3:** Correlation Results for WHO Europe Region

| WHO Europe Region All Countries (n=52) |  |  |  |
| --- | --- | --- | --- |
|  | Government Effectiveness | Regulatory Quality | Control for Corruption |
| GHSI | 0.6 | 0.6 | 0.6 |
| UHC | 0.7 | 0.6 | 0.8 |
| HIS | 0.8 | 0.7 | 0.8 |
| WHO Europe Region Countries excluding Small Countries (n=45) |  |  |  |
|  | Government Effectiveness | Regulatory Quality | Control for Corruption |
| GHSI | 0.6 | 0.6 | 0.7 |
| UHC | 0.7 | 0.6 | 0.8 |
| HIS | 0.8 | 0.7 | 0.8 |
| WHO Europe Region Small Countries (n=7) |  |  |  |
|  | Government Effectiveness | Regulatory Quality | Control for Corruption |
| GHSI | 0.4 | 0.3 | 0.4 |
| UHC | 0.8 | 0.8 | 0.5 |
| HIS | 0.9 | 0.9 | 0.6 |

**Figure 1:**
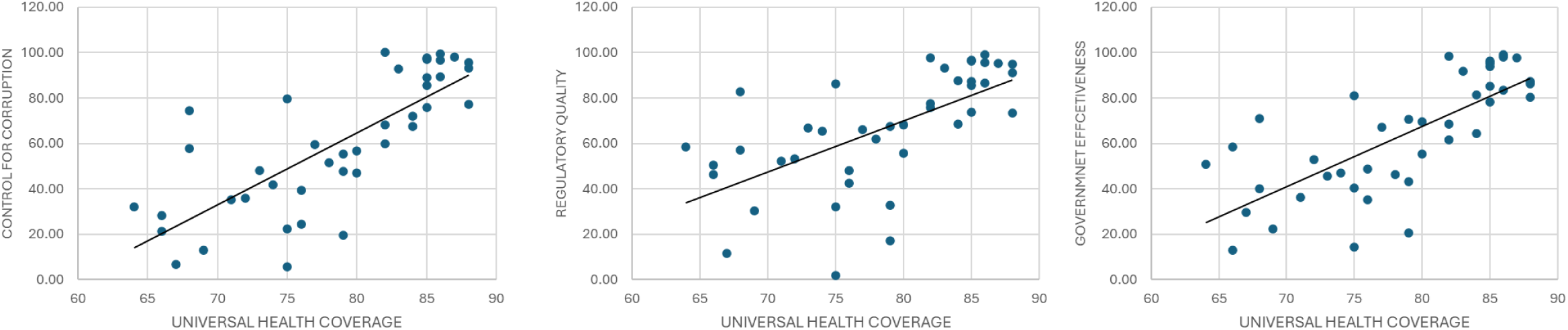
Association between Governance Variables and Universal Health Coverage for 52 WHO Europe Region Countries

In this population good governance is always positively associated with health system outcomes, and government effectiveness is the most important component of the three. Government effectiveness was associated most strongly with the Health Security Index. However, the difference between normal and small countries were significant for both years measured. The results provide evidence that government effectiveness is more important for medium or large countries (0.9) than for smaller countries (0.4). GHSI is also strongly influenced by corruption scores. Similarly, association is much stronger for normal countries (0.9) than small countries (0.4). Although, the association is always positive. Regulatory quality was also strongly associated showing similar patterns. Of the three health related indices, GHSI is the most affected by governance scores.

For UHC is equally highly affected by government effectiveness and corruption scores. A key difference is that government effectiveness seems more important for smaller countries as well. Government effectiveness is equally important for normal and small countries. However, controlling for corruption is much more important for larger countries than smaller ones. Regulatory quality has little less overall impact on UHC (0.5), however, it is still strongly associated. However, it is playing much less role (0.3) for smaller countries.

In this population, HIS is most strongly affected by government effectiveness, the association is equally pronounced for normal and small countries. Corruption is also similarly important for overall HIS, although, much more for normal countries than small ones. Regulatory Quality has a little less effect. However, it is still positive and not to be overlooked (0.5). This also means that regulatory quality is consistently less important for smaller countries as well [it appears] that corruption has less severe impacts in smaller countries.

These WHO Europe Region results reveal that Good governance is always positively associated with health system performance. In addition, Control for corruption and Government effectiveness emerged as relatively more important good governance factors compared to the regulatory quality, even though, the latter has consistently showed a positive association.

The difference between the smaller and larger countries in the WHO Europe region is significant in study of the importance of good governance as an enabler of health system performance. As revealed the small countries have more flexibility and can “*afford*” weaker systems of governance, yet, achieve better results in health system performance. For instance, some smaller, relatively rich countries (the city states) may not have the critical mass of population and education to sensibly engage in sophisticated workforce planning and production however, they may easily access supply from other sources when needed due to their ability to “*purchase*” additional resources and the low numbers involved, thus retaining relatively high UHC scores in the face of region wide access and workforce limitations.

The replication of the WHO Europe country study with the other WHO regions reveals a similar pattern of a positive association between the governance and health system performance variables acting as a confirmation of the WHO Europe results. Table 4 presents the correlation results for the other WHO regions across the governance and health system performance measures. The WHO regional data used for these calculations are included as supplementary information as Tables 5-9 in the Appendix.

**Table 4:**
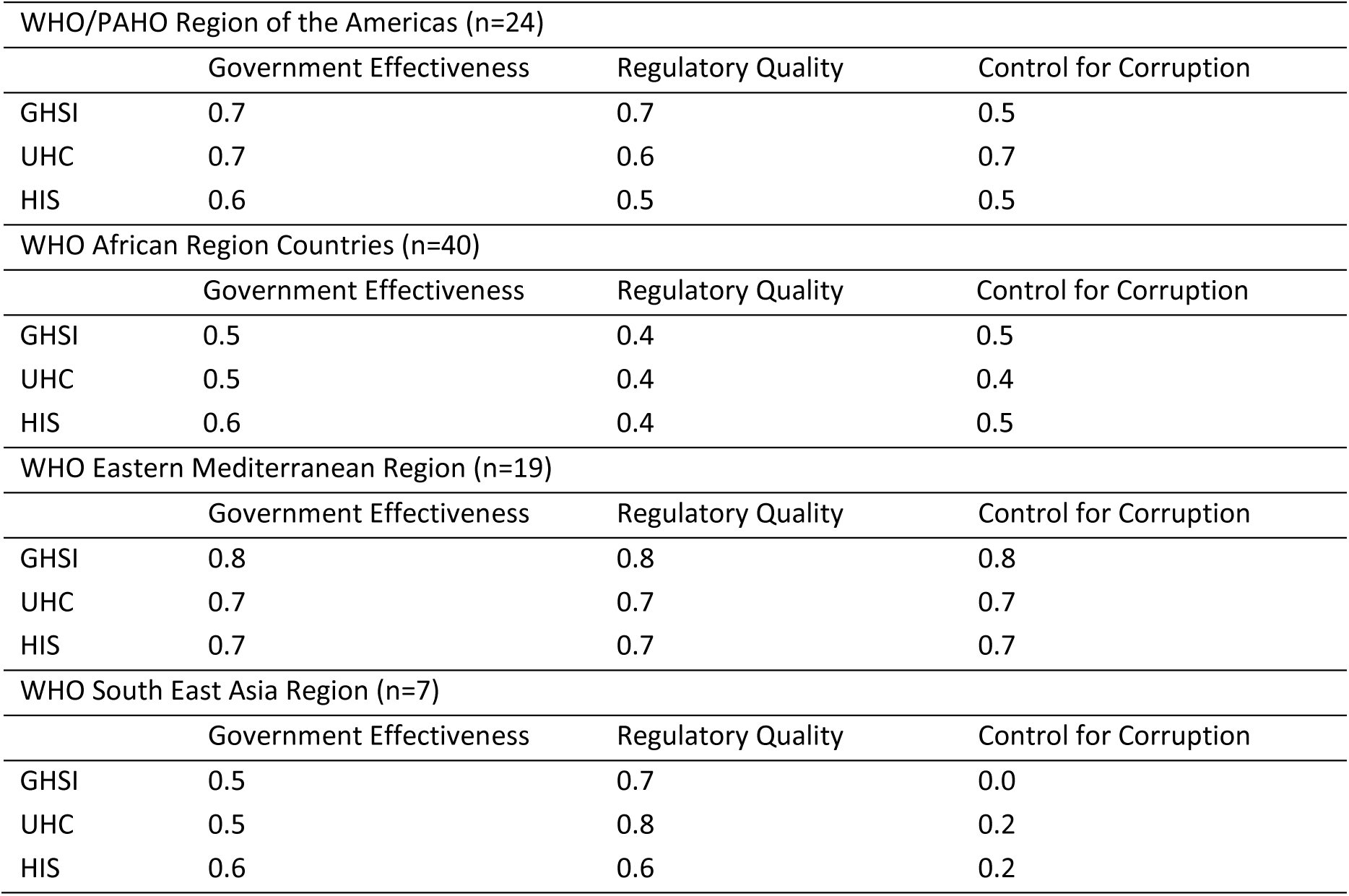
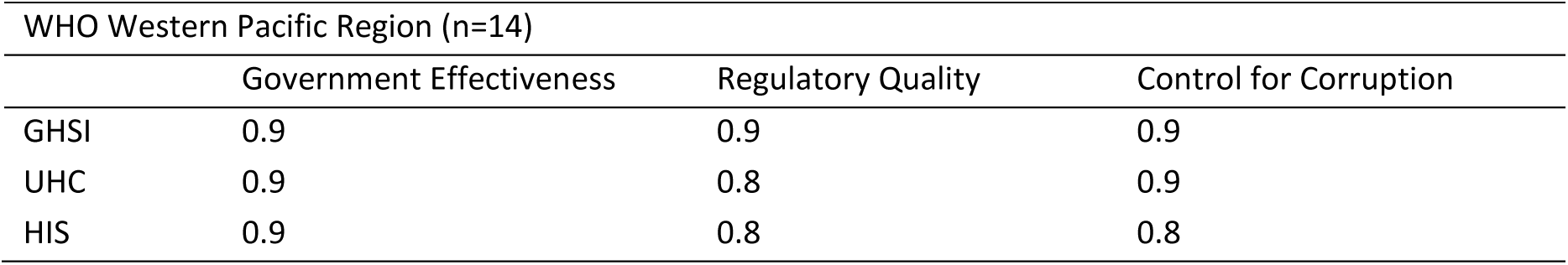
Correlation Results for other WHO Regions

## Discussion

This article aimed to add to the evidence base to bolster the accepted wisdom of the positive association between good governance and health system outcomes by analysing country-level governance and health system indicators. Our findings reveal a positive association between governance and health system performance in the WHO Europe region, which is largely corroborated by the results from the other WHO regions. Although it should be noted that the method used to determine the association does not assure causality between two variables’ relationship [30].

Thus, while these results reveal a positive association between health governance and health system performance, it is certain that more rigorous research is necessary to create additional evidence along the lines of this study’s findings. Henceforth, we suggest further study into potential areas that may deepen our understanding and be able to place greater focus on researching good governance as a likely key enabler of improved health system performance and to better understand regional differences. This is particularly so, for while the regional associations are generally positive some variations exist. The most striking is the association between control for corruption across the performance measures in the South East Asia region sample, which revealed a lower *r* coefficient than found in the other sampled regions.

Though overall, what we find is that an improvement in good governance rankings can lead to the improvement of health index scores. However, we should be clear that good governance itself is influenced by numerous factors, such as political willpower, support, and leadership to mobilize much-needed financial and human resources, but also through well-organized civil service and state institutions, that are able to implement policies and provide services efficiently and effectively. Therefore, one cannot expect quick wins and sustainable improvements in health system performance, especially without adequate improvements in good governance.

Indeed, as indicated previously within a health system framework, governance may be difficult to conceptualize, for on the one hand it can be overarching in nature to reduce the performance dangers of silos and on the other hand it is required to have a localized effect to ensure their optimal functioning [7]. This duality leads to difficulties in the assessment or measurement of governance due to a lack of appropriate indicators [6] or from the difficulty of separating the governance functions from the wider health system and its different system levels [6, 8]. Therefore, even with these results we have generated, we must acknowledge that the effects of each nation’s governance on their health system outcomes may lie within the countries’ specific administrative arrangements, actions and processes, which may or may not be the same nor act at the same levels. For as Batenburg notes, a government’s influence will vary according to a health system’s typology, its planning capacity and operational capabilities [10].

This would indicate for future studies attention should be paid to which particular governance elements, processes or arrangements act to provide the improved health system performances. For governance simply does not mean that everybody agrees [11], rather it is about “how things should be done” juxtaposed by ‘what should be done’, which is the realm of policy making [16, p290]. Thus, as governance is about ensuring that the health system is capable of the overcoming conflicts and divisions between stakeholders to deliver health system outcomes [11], it tends to draw us back to the above paragraph’s issue, which is context.

As Tomoaia-Cotisel et al. report, context is important for most phenomena in health and care, but its defining factors are rarely recorded, included or analysed in research resulting in failed result replication and research translations [32]. Such mechanisms by which governance may influence health system performance have previously been identified as responsiveness to local needs, empowerment of diverse stakeholders, enhanced engagement and strengthened social capital [20], and as Andrews et al., argued, context also needs to be controlled for when developing governance indicators as “countries fall into different categories just as people are in different age groups” [21, p.392]. This indicates that further research on understanding the types and effects of governance arrangements, key interactions between governance stakeholders and the levels at which they operate will cast more light on the identification of these factors of health governance’s influence on health system outcomes. Some of the governance components, while possibly exercising different effects have already been classified in the literature such as (i) strategic vision and policymaking, (ii) participation/partnerships/collaboration, (iii) transparent, data-driven and evidence-based decisions, and (iv) legislation and regulation towards public health goals [7]. By taking such a focus the identification of countries that seem to have governance problems may be enabled, which then can then be followed up with more detailed analyses of what the problems are where they lie [21]

This importance of context is reinforced by the results in terms of the small country sub-sample differences from the rest of the WHO Europe sample. Here associations between particular variables were found to be different, highlighted by the result for government effectiveness’ importance for larger countries over that for smaller countries. These differences are also repeated across other variables illustrating how context is important and that from a more specified study, and understanding of how the essential contextual differences between small state health governance and larger states matter. Nevertheless, these data provide a starting point for an investigation of governance in small states to further understand how they may mobilize governance for improved health system performances.

## Conclusion

Overall, our results show that it is evident that good governance has some impact on health system performance. Good governance is influenced by numerous factors, but most importantly, by political willpower, support, and leadership, especially from the highest levels of government. Thus, our results also suggest that countries with better governance and poor planning may be expected to achieve better health outcomes than countries with poor governance but better planning, and there is unlikely to be a quick win in improving health outcomes without improvements in good governance. In addition, we found that for some smaller, relatively rich countries (city states) there are different strengths of associations than for larger European countries. This reinforces the contextual nature of governance and as such we reiterate the cautions contained in the literature and encourage the recognition of context for future research in this field.

## Data Availability

All the data used for this article are included in the manuscript's text.

## Appendix

**Table 5:** Data from the WHO/PAHO Region of the Americas

| Countries | Global Health Security Index (2021) | UHC Service Coverage Index (WHO, 2021) | Health Index Score (Statista, 2023) | Government Effectiveness (WB, 2021) | Regulatory Quality (WB, 2021) | Control for Corruption (WB, 2021) |
| --- | --- | --- | --- | --- | --- | --- |
| Argentina | 54.4 | 79 | 74.5 | 37.14 | 30.00 | 38.10 |
| Belize | 29.7 | 68 | 70.6 | 32.86 | 36.67 | 44.29 |
| Bolivia | 29.9 | 65 | 66 | 23.81 | 10.95 | 20.48 |
| Brazil | 51.2 | 80 | 71.7 | 33.81 | 47.14 | 34.29 |
| Canada | 69.8 | 91 | 78.9 | 94.29 | 94.29 | 92.38 |
| Chile | 56.2 | 82 | 76.1 | 70.00 | 78.57 | 80.95 |
| Colombia | 53.2 | 80 | 77.8 | 50.48 | 60.00 | 42.38 |
| Costa Rica | 40.8 | 81 | 79.1 | 59.52 | 67.14 | 66.67 |
| Cuba | 30.5 | 83 | 79.5 | 41.90 | 6.19 | 54.29 |
| Dominican Republic | 34.5 | 77 | 70.6 | 51.90 | 56.19 | 30.95 |
| Ecuador | 50.8 | 77 | 71.8 | 42.38 | 25.71 | 30.00 |
| El Salvador | 40.8 | 78 | 69.3 | 38.10 | 39.52 | 32.86 |
| Guatemala | 29.1 | 59 | 68.3 | 32.86 | 40.48 | 11.90 |
| Guyana | 30.8 | 76 | 62.5 | 40.95 | 33.81 | 48.57 |
| Honduras | 26.2 | 64 | 69 | 20.00 | 33.33 | 14.76 |
| Mexico | 57 | 75 | 73.1 | 37.62 | 44.29 | 17.62 |
| Nicaragua | 36.3 | 70 | 72.1 | 17.62 | 19.52 | 9.52 |
| Panama | 53.5 | 78 | 75.1 | 57.14 | 59.05 | 30.48 |
| Paraguay | 40.3 | 72 | 70.8 | 28.10 | 44.76 | 16.67 |
| Peru | 54.9 | 71 | 75.8 | 39.52 | 55.24 | 28.57 |
| Suriname | 35 | 63 | 60.8 | 26.67 | 25.24 | 39.05 |
| United States of America | 75.9 | 86 | 73.3 | 87.62 | 90.48 | 82.86 |
| Uruguay | 40.3 | 82 | 78 | 75.24 | 72.86 | 91.43 |
| Venezuela | 20.9 | 75 | 69.6 | 1.90 | 0.95 | 1.90 |

Note: Data for Antigua and Barbuda, Bahamas, Barbados, Dominica, Grenada, Haiti, Jamaica, Sait Kitts and Nevis, Saint Lucia, Saint Vincent and the Grenadines, Trinidad and Tobago are not included.

**Table 6:** Data from the WHO African Region

| Countries | Global Health Security Index (2021) | UHC Service Coverage Index (WHO, 2021) | Health Index Score (Statista, 2023) | Government Effectiveness (WB, 2021) | Regulatory Quality (WB, 2021) | Control for Corruption (WB, 2021) |
| --- | --- | --- | --- | --- | --- | --- |
| Algeria | 26.2 | 74 | 73.2 | 28.57 | 10.00 | 29.52 |
| Angola | 29.1 | 37 | 49.9 | 12.38 | 27.14 | 29.05 |
| Benin | 25.4 | 38 | 53.5 | 42.86 | 36.19 | 49.05 |
| Botswana | 33.6 | 55 | 59.1 | 63.33 | 71.90 | 74.76 |
| Burkina Faso | 29.8 | 40 | 58.7 | 23.33 | 35.24 | 50.48 |
| Burundi | 22.1 | 41 | 58.2 | 8.57 | 13.33 | 2.86 |
| Cameroon | 28.6 | 44 | 51 | 16.19 | 16.67 | 13.33 |
| Central African Republic | 18.6 | 32 | 32 | 4.76 | 5.24 | 10.95 |
| Chad | 23.9 | 29 | 39.2 | 6.67 | 10.48 | 4.76 |
| Congo (Brazzaville) | 26.3 | 41 | 54.1 | 9.05 | 9.52 | 6.19 |
| Congo (Democratic Republic) | 26.1 | 42 | 50.9 | 3.33 | 6.67 | 4.29 |
| Côte d'Ivoire | 31.2 | 43 | 51.8 | 32.38 | 43.33 | 40.48 |
| Equatorial Guinea | 17.4 | 46 | 50.4 | 11.43 | 3.81 | 3.33 |
| Eritrea | 21.4 | 45 | 57.5 | 4.29 | 0.48 | 9.05 |
| eSwatini | 29.3 | 56 | 48.4 | 25.71 | 31.43 | 27.14 |
| Ethiopia | 37.8 | 35 | 60.9 | 29.05 | 16.19 | 37.62 |
| Gabon | 21.8 | 49 | 56.1 | 19.52 | 21.90 | 20.95 |
| Gambia | 28.7 | 46 | 56.2 | 27.14 | 18.57 | 40.95 |
| Ghana | 34.3 | 48 | 63.1 | 44.76 | 45.71 | 50.00 |
| Guinea | 26.8 | 40 | 48.7 | 14.76 | 13.81 | 17.14 |
| Guinea-Bissau | 21.4 | 37 | 50.9 | 7.14 | 8.57 | 7.14 |
| Kenya | 38.8 | 53 | 65.2 | 38.57 | 35.71 | 40.00 |
| Lesotho | 30.9 | 53 | 41.3 | 15.24 | 24.76 | 43.33 |
| Liberia | 35.7 | 45 | 48.6 | 8.10 | 14.76 | 19.05 |
| Madagascar | 30.4 | 35 | 54.1 | 13.81 | 20.95 | 18.57 |
| Malawi | 28.5 | 48 | 61 | 21.43 | 22.38 | 44.76 |
| Mauritania | 26.2 | 40 | 56.7 | 22.86 | 12.86 | 21.90 |
| Mozambique | 30.4 | 44 | 52.8 | 25.24 | 22.86 | 22.86 |
| Namibia | 30.3 | 63 | 60.6 | 54.76 | 51.43 | 61.43 |
| Niger | 28.7 | 35 | 54.6 | 30.48 | 24.29 | 31.43 |
| Nigeria | 38 | 38 | 50.1 | 13.33 | 15.71 | 14.29 |
| Rwanda | 33.1 | 49 | 64.6 | 60.00 | 53.81 | 70.00 |
| Senegal | 32.8 | 50 | 64.4 | 53.81 | 40.95 | 57.14 |
| Sierra Leone | 32.7 | 41 | 48.3 | 11.90 | 14.29 | 36.19 |
| South Africa | 45.8 | 71 | 59.9 | 50.00 | 49.52 | 53.81 |
| Tanzania | 31.3 | 43 | 60.1 | 30.00 | 29.05 | 26.19 |
| Togo | 27.8 | 44 | 55.1 | 26.19 | 28.10 | 27.62 |
| Uganda | 36.5 | 49 | 58.5 | 31.43 | 34.76 | 16.19 |
| Zambia | 26.5 | 56 | 57.2 | 18.10 | 32.38 | 25.71 |
| Zimbabwe | 32.4 | 55 | 55.5 | 10.48 | 7.14 | 10.00 |

Note: Data for Cabo Verde, Comoros, Mali, Mauritius, Sao Tome and Principe, Seychelles are not included.

**Table 7:**
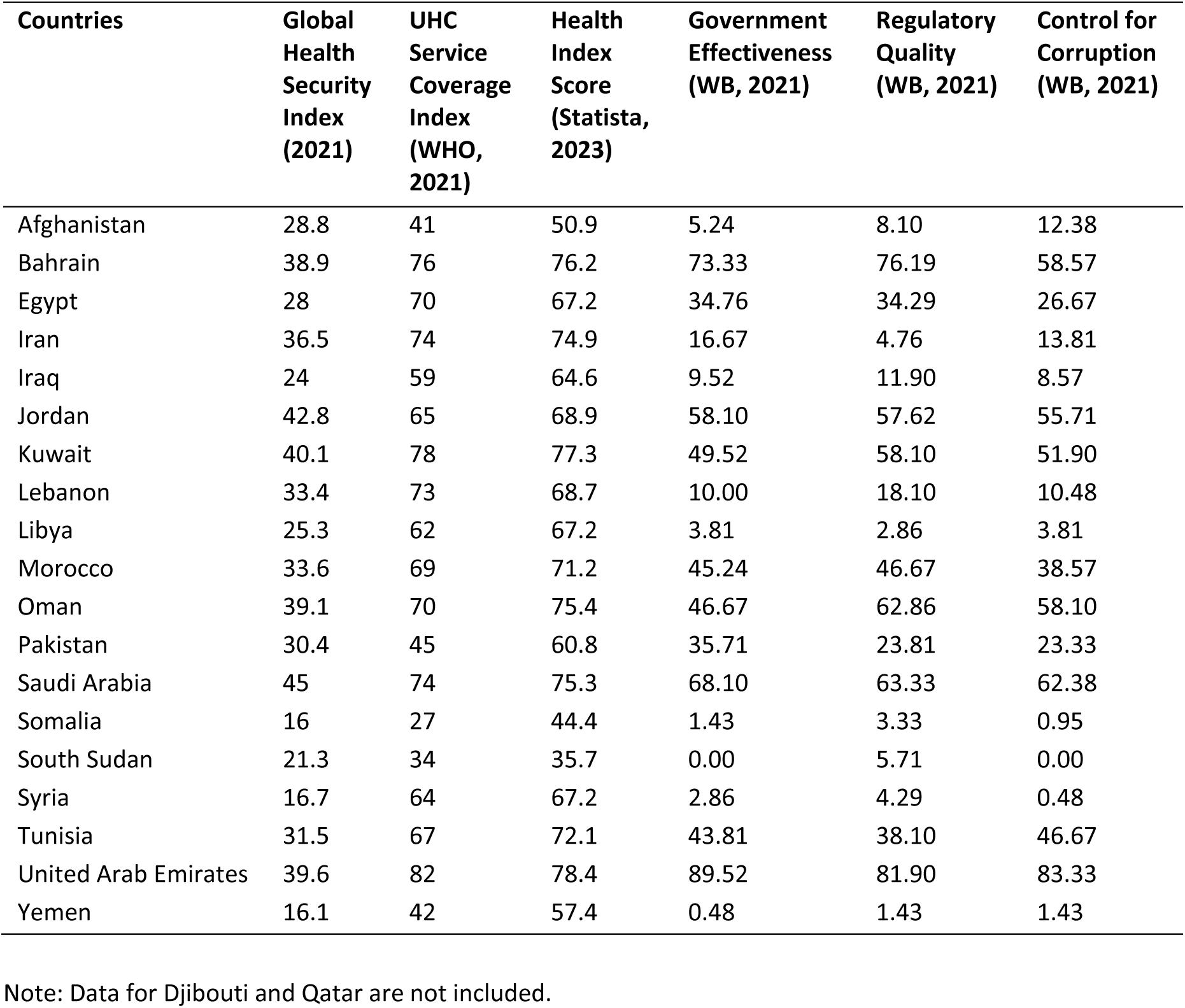
Data from the WHO Eastern Mediterranean Region

**Table 8:** Data from the WHO South East Asia Region

| **Countries** | **Global Health Security Index (2021)** | **UHC Service Coverage Index (WHO, 2021)** | **Health Index Score (Statista, 2023)** | **Government Effectiveness (WB, 2021)** | **Regulatory Quality (WB, 2021)** | **Control for Corruption (WB, 2021)** |
| --- | --- | --- | --- | --- | --- | --- |
| Bangladesh | 35.5 | 52 | 67.4 | 27.62 | 20.48 | 18.10 |
| Bhutan | 39.8 | 60 |  | 74.29 | 37.14 | 90.48 |
| India | 42.8 | 63 | 66.2 | 60.95 | 49.05 | 45.24 |
| Indonesia | 50.4 | 55 | 71.1 | 64.76 | 60.95 | 36.67 |
| Myanmar | 38.3 | 52 | 66.7 | 7.62 | 12.38 | 15.71 |
| Nepal | 34 | 54 | 63.9 | 15.71 | 28.57 | 32.38 |
| Thailand | 68.2 | 82 | 78.9 | 59.05 | 56.67 | 34.76 |

Note: Data for the Democratic People's Republic of Korea, the Maldives, Sri Lanka and Timor-Leste are not included.

**Table 9:** Data from the WHO Western Pacific Region

| **Countries** | **Global Health Security Index (2021)** | **UHC Service Coverage Index (WHO, 2021)** | **Health Index Score (Statista, 2023)** | **Government Effectiveness (WB, 2021)** | **Regulatory Quality (WB, 2021)** | **Control for Corruption (WB, 2021)** |
| --- | --- | --- | --- | --- | --- | --- |
| Australia | 71.1 | 87 | 80.4 | 92.86 | 98.57 | 94.76 |
| Brunei | 43.5 | 78 |  | 90.48 | 81.43 | 85.24 |
| Cambodia | 31.1 | 58 | 70.6 | 34.29 | 26.67 | 11.43 |
| China | 47.5 | 81 | 83.1 | 75.71 | 40.00 | 56.19 |
| Japan | 60.5 | 83 | 86.5 | 89.05 | 90.00 | 90.95 |
| Laos | 34.8 | 52 | 66.4 | 30.95 | 17.62 | 15.24 |
| Malaysia | 56.4 | 76 | 77.3 | 80.00 | 72.38 | 59.05 |
| Mongolia | 41 | 65 | 66.7 | 33.33 | 43.81 | 33.33 |
| New Zealand | 62.5 | 85 | 79.8 | 88.10 | 98.10 | 99.05 |
| Papua New Guinea | 25 | 30 | 52.9 | 17.14 | 21.43 | 24.76 |
| Philippines | 45.7 | 58 | 70.1 | 55.71 | 54.76 | 33.81 |
| Republic of Korea | 65.4 | 89 | 84.8 | 90.00 | 83.33 | 76.67 |
| Singapore | 57.4 | 89 | 86.9 | 100.00 | 100.00 | 98.57 |
| Viet Nam | 42.9 | 68 | 77 | 60.48 | 37.62 | 45.71 |

Note: Data for the Cook Islands, Fiji, Kiribati, Marshall Islands, Federated states of Micronesia, Nauru, Niue, Palau, Samoa, Solomon Islands, Tonga, Tuvalu and Vanuatu are not included.

## Notes

### Competing Interest Statement

The authors have declared no competing interest.

### Funding Statement

This study did not receive any external funding.

## References

1. Rathnayake D, Clarke M, Jayasinghe VI. Health system performance and health system preparedness for the post-pandemic impact of COVID-19: A review. International Journal of Healthcare Management. 2020;14(1):250–254. 10.1080/20479700.2020.1836732

2. World Health Organization. Everybody’s Business -- Strengthening Health Systems to Improve Health Outcomes: WHO’s Framework for Action. World Health Organization; 2007. 9789241596077

3. World Health Organisation. Promoting health through good governance. www.who.int. Published 2024. https://www.who.int/activities/promoting-health-through-good-governance Accessed February 22, 2024

4. Martineau T, Ozano K, Raven J, et al. Improving health workforce governance: the role of multi-stakeholder coordination mechanisms and human resources for health units in ministries of health. Human Resources for Health. 2022;20(47). 10.1186/s12960-022-00742-z

5. Debie A, Khatri RB, Assefa Y. Successes and challenges of health systems governance towards universal health coverage and global health security: a narrative review and synthesis of the literature. Health Research Policy and Systems. 2022;20(50). 10.1186/s12961-022-00858-7

6. Pyone T, Smith H, van den Broek N. Frameworks to assess health systems governance: a systematic review. Health Policy and Planning. 2017;32(5):710–722. 10.1093/heapol/czx007

7. Rajan D, Koch K, Rohrer K, Soucat A. Governance. In: Papanicolas I, Rajan D, Karanikolos M, Soucat A, Figueras J, eds. Health System Performance Assessment: A Framework for Policy Analysis. (Health Policy Series, No. 57). World Health Organization; 2022:43–77. https://iris.who.int/handle/10665/352686

8. Mikkelsen-Lopez I, Wyss K, de Savigny D. An approach to addressing governance from a health system framework perspective. BMC International Health and Human Rights. 2011;11(13). 10.1186/1472-698x-11-13

9. Hastings SE, Armitage GD, Mallinson S, Jackson K, Suter E. Exploring the relationship between governance mechanisms in healthcare and health workforce outcomes: a systematic review. BMC Health Services Research. 2014;14(479). 10.1186/1472-6963-14-479

10. Batenburg R. Health workforce planning in Europe: Creating learning country clusters. Health Policy. 2015;119(12):1537–1544. 10.1016/j.healthpol.2015.10.001

11. Greer SL, Wismar M, Figueras J, McKee C. Governance: a framework. In: Greer SL, Wismar M, Figueras J, eds. Strengthening Health System Governance: Better Policies, Stronger Performance. Open University Press; 2016:27–46. https://eurohealthobservatory.who.int/publications/m/strengthening-health-system-governance-better-policies-stronger-performance Accessed February 23, 2024.

12. Gostin LO, Mok EA. Grand challenges in global health governance. British Medical Bulletin. 2009;90(1):7–18. 10.1093/bmb/ldp014

13. Zhang H. Challenges and Approaches of the Global Governance of Public Health Under COVID-19. Frontiers in Public Health. 2021;9. 10.3389/fpubh.2021.727214

14. Kickbusch I, Gleicher D. *Governance for Health in the 21st Century*. World Health Organization, Regional Office for Europe; 2012.

15. Azimi MN, Rahman MM, Nghiem S. A global perspective on the governance-health nexus. BMC Health Services Research. 2023;23(1235). 10.1186/s12913-023-10261-9

16. Travis P, Egger D, Davies P, Mechbal A. Towards better stewardship: concepts and critical issues. In: Murray CJ, Evans DB, eds. Health Systems Performance Assessment: Methods, Debate and Empiricism. WHO Regional Office for Europe; 2003:289–300. https://www.who.int/publications/i/item/9241562455 Accessed February 22, 2024

17. Papanicolas I, Karanikolos M, Figueras J, Rajan D. Working towards a common approach: the HSPA Framework for UHC. In: Papanicolas I, Rajan D, Karanikolos M, Soucat A, Figueras J, eds. Health System Performance Assessment: A Framework for Policy Analysis. World Health Organization; 2022:26–42. https://iris.who.int/handle/10665/352686 Accessed February 22, 2024

18. Barbazza E, Langins M, Kluge H, Tello J. Health workforce governance: Processes, tools and actors towards a competent workforce for integrated health services delivery. Health Policy. 2015;119(12):1645–1654. 10.1016/j.healthpol.2015.09.009

19. Masefield SC, Msosa A, Grugel J. Challenges to effective governance in developing health systems: a qualitative study in Malawi. European Journal of Public Health. 2020;30(Supplement_5). 10.1093/eurpub/ckaa166.504

20. Ciccone DK, Vian T, Maurer L, Bradley EH. Linking governance mechanisms to health outcomes: A review of the literature in low- and middle-income countries. Social Science & Medicine. 2014;117:86–95. 10.1016/j.socscimed.2014.07.010

21. Andrews M, Hay R, Myers J. Can Governance Indicators Make Sense? Towards a New Approach to Sector-Specific Measures of Governance. Oxford Development Studies. 2010;38(4):391–410. 10.1080/13600818.2010.524696

22. Koller T, Clarke D, Vian T. Promoting anti-corruption, transparency and accountability to achieve universal health coverage. Global Health Action. 2020;13(sup1):1700660. 10.1080/16549716.2019.1700660

23. World Bank. Worldwide Governance Indicators. World Bank. Published 2023. https://www.worldbank.org/en/publication/worldwide-governance-indicators Accessed February 24, 2024.

24. World Health Organization. Indicator Metadata Registry Details. www.who.int. Published 2023. https://www.who.int/data/gho/indicator-metadata-registry/imr-details/4834 Accessed February 24, 2024.

25. Legatum Institute. Rankings: Legatum Prosperity Index 2018. Legatum Prosperity Index 2018. Published November 27, 2018. https://www.prosperity.com/rankings Accessed February 24, 2024.

26. Reid M, Gupta R, Roberts G, Goosby E, Wesson P. Achieving Universal Health Coverage (UHC): Dominance analysis across 183 countries highlights importance of strengthening health workforce. Mathur MR, ed. PLOS ONE. 2020;15(3):e0229666. 10.1371/journal.pone.0229666

27. Leegwater A, Wong W, Avila C. A concise, health service coverage index for monitoring progress towards universal health coverage. BMC Health Services Research. 2015;15(230). 10.1186/s12913-015-0859-3

28. The 2021 Global Health Security Index. GHS Index. Published 2021. https://ghsindex.org/ Accessed February 24, 2024.

29. Sedgwick P. Pearson’s correlation coefficient. BMJ. 2012;345:e4483–e4483. 10.1136/bmj.e4483

30. Schober P, Boer C, Schwarte LA. Correlation coefficients: Appropriate Use and Interpretation. Anesthesia & Analgesia. 2018;126(5):1763–1768. 10.1213/ANE.0000000000002864

31. Tomoaia-Cotisel A, Scammon DL, Waitzman NJ, et al. Context Matters: The Experience of 14 Research Teams in Systematically Reporting Contextual Factors Important for Practice Change. The Annals of Family Medicine. 2013;11(Suppl_1):S115–S123. 10.1370/afm.1549

32. World Health Organization. Roadmap towards Better Health in Small Countries in the WHO European Region, 2022–2025. World Health Organization Regional Office for Europe; 2022:1–16.. https://www.who.int/europe/publications/i/item/WHO-EURO-2022-5484-45249-64713 Accessed February 24, 2024

33. World Health Organization. Countries overview | World Health Organization. https://www.who.int/countries/ Accessed February 22, 2024.

